# Digital Cognitive Behavioral Therapy for Cardiac Anxiety Following Acute Coronary Syndrome: Protocol for a Randomised Controlled Trial Comparing CBT to a Digital Lifestyle Intervention

**DOI:** 10.1101/2025.06.11.25329458

**Authors:** Amanda Johnsson, Brjánn Ljótsson, Frieder Braunschweig, Linda G Mellbin, Josefin Särnholm

## Abstract

**Introduction:** Cardiac anxiety is common following acute coronary syndrome (ACS) and is characterised by fear of recurrence, heightened attention to cardiac sensations, and avoidance of cardiac-related activities in daily life. It is associated with depression, reduced quality of life (QoL), and an adverse cardiac prognosis. We have developed a digital cognitive behavioural therapy protocol for cardiac anxiety (CA-CBT) post ACS, previously evaluated in clinical trials including one randomized controlled trial (RCT), in which the intervention was compared to usual care. This protocol article describes a follow-up RCT, designed to further evaluate the efficacy of CA-CBT compared to an active control receiving a digital cardiac life-style intervention (CLI).

**Method and analysis:** Participants with ACS (STEMI/Non-STEMI /Unstable angina ≥ 6 months prior) and elevated cardiac anxiety (Cardiac Anxiety Questionnaire; CAQ≥18 and as per clinical interviews) (n=176) are randomised 1:1 to 8 weeks of CA-CBT or CLI. Primary outcome is change in cardiac anxiety, measured by the CAQ, from pre-to post-intervention. Analyses will be conducted according to the ‘intention-to-treat’ principle, using hierarchical linear mixed-effects model, with random intercepts, and include ten weekly assessments collected during the treatment period. Secondary outcomes include disease-specific Quality of Life (Heart-Qol), depression (PHQ-9) insomnia (ISI), in addition to both self-rated and accelerometer-measured physical activity. Secondary outcomes will be analysed using similar statistical methods.

**Ethics and dissemination:** The study was approved by the Swedish Ethical Review Authority (Dnr 2023-07605-01), and the first patient enrolled March 7 2024. Recruitment is ongoing and is expected to conclude in the latter half of 2026. All participants receive information about the study and provide informed consent in accordance with ethical guidelines before inclusion. The results will be analysed at group level, and trial outcomes will be published in a peer-reviewed scientific journal, regardless of results.

**Trial registration number:** Registered at ClinicalTrials.gov, ID: NCT06298864

**Strengths and limitations:**

- To our knowledge, this is the first exposure-based CBT intervention specifically targeting cardiac anxiety following ACS, addressing a modifiable psychological risk factor with relevance for secondary prevention.
- The use of an active control group helps account for non-specific effects. However, overlapping components, such as behavioural strategies to increase physical activity, between the CBT and lifestyle interventions poses a risk of diluting between-group differences.
- The use of objective physical activity measures, assessed via accelerometry, alongside well-validated self-report instruments, strengthens the validity of the outcome measurements.
- The study procedures include a multidisciplinary assessment and collaborative treatment delivery to ensure patient safety.
- Manual-based treatment delivery with therapist support ensures minimal therapist drift while maintaining an individualised approach. Using the same therapist across both study arms helps reduce the risk of therapist-related confounding effects.

## BACKGROUND AND RATIONALE

Acute coronary syndrome (ACS), encompassing unstable angina (UA) and myocardial infarction (MI), is an acute cardiovascular event (1) and a leading global cause of mortality and health-related losses (2). Psychological distress, such as anxiety and depression, is common in patients after an ACS (3). A disease specific form of psychological distress is cardiac anxiety, due to its specific focus on cardiac-related concerns. Cardiac anxiety is defined as an excessive fear of recurrence, heightened attention to cardiac-related sensations (e.g. palpitations, shortness of breath) and cardiac-related related avoidance behaviours (e.g. physical activity, leaving the house) (4). Following ACS, approximately one third of patients develops this type of anxiety (5). Cardiac anxiety, and in particular its avoidance component, has been linked to the development of depressive symptoms (5), poorer quality of life (QoL) (6), and an adverse cardiac prognosis (7). Cardiac anxiety has also been identified as a barrier to engage in health behaviors, including physical activity and participation in cardiac rehabilitation (6, 8). For many patients, cardiac-related sensations, such as palpitations or shortness of breath, have been paired with fear via interoceptive fear conditioning, and elicit a fear response (9) which in turn drives avoidance behaviour. Cardiac anxiety may also extend to a broad range of situations perceived as unsafe, such as leaving the house, travelling or walking alone, further limiting the patient from engaging in previously enjoyed activities. (10) Despite its clinical relevance, structured psychological interventions specifically targeting cardiac anxiety remain underutilised in routine post-ACS care (11).

Cardiac rehabilitation, including lifestyle modification, is a key component in regaining QoL following ACS (1, 12). Interventions focusing on lifestyle modification and health behaviours, including digitally delivered cardiac-rehabilitation, have proven effective in restoring health and mitigating the risk of recurrent cardiovascular events (13, 14). While guidelines for supporting health behaviours following ACS are well established (1, 12), mental health remains largely under-addressed and undertreated in this population highlighting a gap in both available and implemented treatment options (11).

Cognitive behavioural therapy (CBT) is an evidence-based form of psychological intervention. Studies on CBT for patients with cardiovascular disease, including those with ACS, have demonstrated reductions of anxiety, depression (15, 16), as well as decreased risk of readmissions (17), and recurrent cardiac events (18). Furthermore, the digital deliverance for such interventions has proven effective (19). While psychological interventions for coronary heart disease have traditionally focused on addressing general anxiety and depression, a recent Cochrane review underscores the importance of developing approaches more adapted to the patient’s psychological clinical presentation (15). Exposure-based CBT targets the cycle of avoidance and fear of cardiac-related sensations, situations and disability by gradually confronting the stimuli and situations associated with cardiac-related fear and distress (20). Focusing specifically on the reduction of avoidance behaviours through exposure may in fact be a key to reduce anxiety following ACS, and improve QoL.

Särnholm et al. (20–22) have previously developed an exposure-based digital CBT intervention for cardiac anxiety in patients with atrial fibrillation (AF-CBT), which resulted in large reduction in cardiac anxiety and improvement in disease-specific QoL. In the present research project, we further adapted and tailored the exposure-based digital CBT intervention to patients with elevated cardiac anxiety post ASC in the cardiac anxiety CBT (CA-CBT). The intervention proved feasible and clinically promising in two pilot studies (23) and a randomised controlled trial (RCT). The RCT comparedCA-CBT to usual care (Johnsson et al., manuscript in preparation; ClinicalTrials: NCT05580718) and demonstrated a substantial reduction in cardiac anxiety and an improvement in QoL, in favor of the CA-CBT group.

As our previous trial compared CBT to usual care, uncertainty remains regarding the specific efficacy of CBT beyond general therapeutic attention, structure, and support. To address this limitation, the present study includes an active attention control: a digital cardiac lifestyle intervention matched in duration, format, and delivery mode. This design enables the isolation of the specific effects of CBT techniques, such as exposure, while controlling for non-specific factors including therapist contact and expectancy effects (24). By comparing CBT-ACS to a credible, structured comparator, the study enhances internal validity and more closely reflects real-world clinical contexts—where patients are often offered some form of lifestyle support—allowing us to also determine whether CBT provides added value. This protocol article outlines the methods and procedures of the study.

### Objectives

The objective of this RCT is to further evaluate the CA-CBT compared to an active attention control condition receiving a digital cardiac lifestyle intervention (CLI) incorporating behavioural change strategies.

We hypothesise that exposure-based digital CBT targeting cardiac anxiety following ACS is effective in reducing cardiac anxiety and depressive symptoms, while improving disease-specific QoL and physical activity at post-intervention. We anticipate that the improvements will be maintained at six and twelve-months follow-up.

## METHODS

### Study design

This is a two-arm, parallel-group RCT and that aim to include 176 participants, who are randomised 1:1 to CA-CBT (N=88) or CLI (N=88) over an eight week period. The design aim to isolate specific treatment effects and control for unspecific factors, such as attention, therapeutic alliance and expectancy of improvement (24). In the absence of a gold-standard treatment, we selected an active attention control with content that is both recommended and credible for the target population (12), yet distinct in the hypothesised active element of exposure. The active control (CLI) was designed to match CBT in terms of deliverance, structure and duration. Participants remain blinded to group allocation throughout the study and are not informed about the content or nature of the intervention to which they are not assigned. This protocol has been developed in accordance with the Standard Protocol Items: Recommendations for Interventional Trials (SPIRIT) 2013 guidelines. A completed SPIRIT checklist is provided as Supplementary Material.

### Study setting

Participants are recruited nationwide in Sweden through online self-enrolment, as further outlined below. The assessments are performed remotely, data-collection and study participation are conducted digitally and the trial is based at Karolinska University Hospital, Stockholm, Sweden. All materials are in Swedish.

### Eligibility criteria

Inclusion criteria: (A) ACS ≥ 6 months before assessment (type 1 MI STEMI/NSTEMI or UA); (B) Age 18 and older; (C) (Cardiac Anxiety Questionnaire; CAQ: ≥18); (D) Clinically significant cardiac anxiety that leads to distress and/or interferes with daily life as per clinical interview; (D) Able to read and write in Swedish.

Exclusion criteria:(F) Heart failure New York heart Association class IV (25) or ejection fraction ≤30%; (G) Significant valvular disease; (H) Planned coronary artery bypass surgery or percutaneous interventions; (I) Any medical restriction to physical exercise; (J) Severe medical illness or an acute health threatening disease (e.g., cancer); (K) Grade 3 hypertension (i.e., blood pressure ≥ 180 systolic and/or 110 diastolic); (L) Severe mental illness requiring other primary intervention, psychiatric hospitalisation or elevated risk of suicide; (M) Alcohol or substance use disorder that would impede ability to complete study protocol; (N) Ongoing psychological treatment.

### Recruitment

Participants are recruited via self-referral from all of Sweden through advertisements in social media, daily press, and information directed to cardiology clinics, medical clinics and primary care units. Furthermore, medical records from Karolinska University Hospital, Stockholm, are used to identify potential participants who have experienced an ACS, and written information about the study is sent to them by mail.

Interested applicants complete an online screening and provide informed consent through a secure web-based platform and server (BASS). The online screening includes the Cardiac Anxiety Questionnaire (CAQ) (8), HeartQoL(30) and Patient Health Questionnaire-9 (PHQ-9) (33). To screen for alcohol dependency, the screening also includes the Alcohol Use Disorders Identification Test (AUDIT)(45). In addition to this, it also includes questions on demographic information and medical history.

Following the screening, a chart review is conducted by the cardiac study nurse to assess ACS classification, comorbid conditions, medical history and key cardiac parameters, including recent blood pressure, lipid status, ECG, echocardiogram results (if available), and current medications. Thereafter, the nurse proceeds with a telephone assessment. During this assessment, the cardiac study nurse confirms medical history and collect key cardiac parameters. Eligible applicants then undergo a telephone-based psychological assessment by a study psychologist, to screen for clinically significant cardiac anxiety and exclude patients with severe psychiatric disorder, severe depression, risk of suicide and/or alcohol dependency. The medical assessments and the cardiac parameters from the medical records for eligible applicants are then reviewed by the study cardiologist, to determine whether the participant meets the eligibility criteria. Figure 1 illustrates the participant flow through the study.

**Figure 1.**
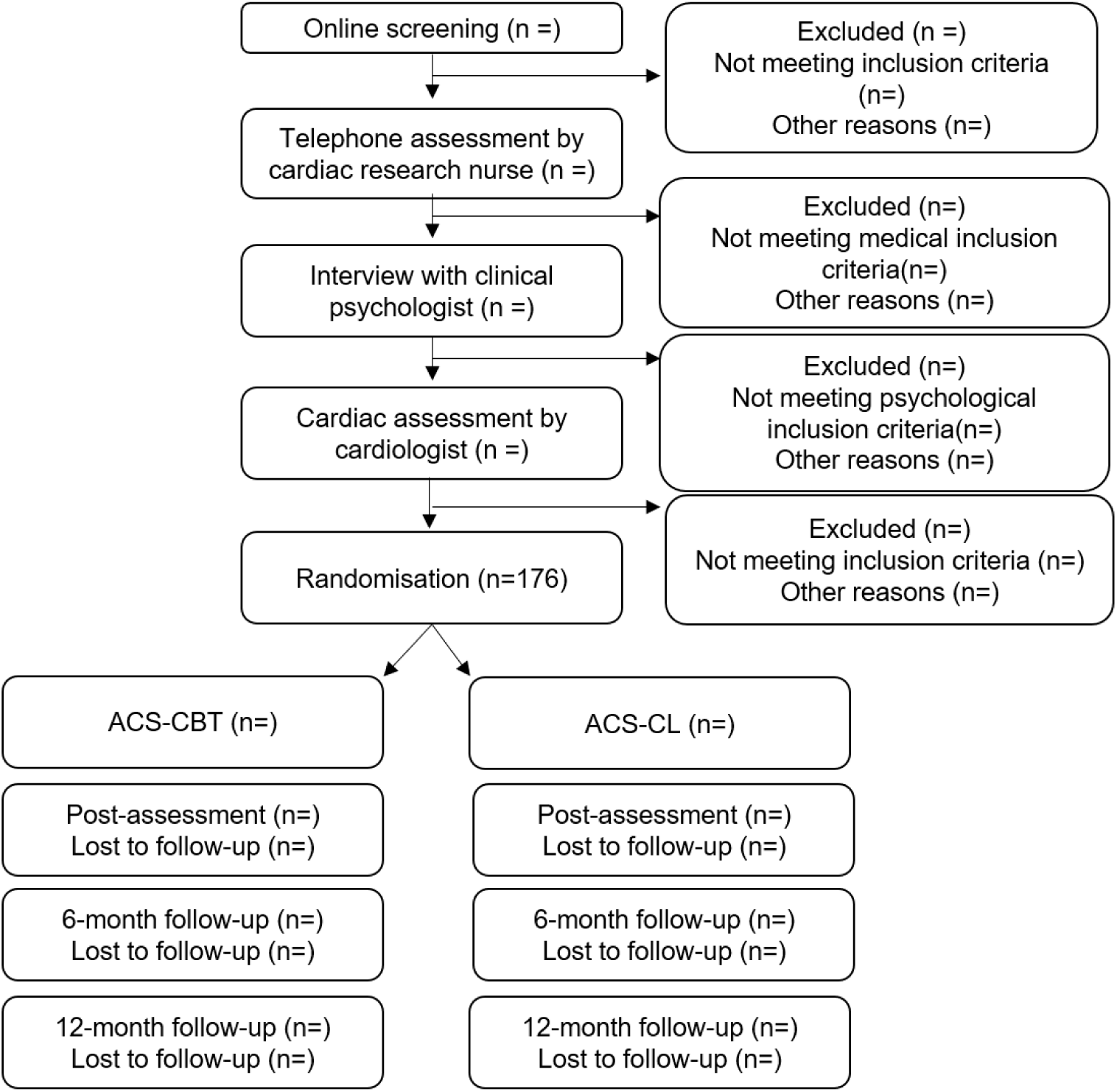
Flow chart. Participants flow through the study.

### Randomisation and allocation

Participants are randomised by an external party, not connected to the study, using a true random number service (www.random.org). The randomisation follows a 1:1 ratio, assigning participants to either CA-CBT or CLI. Randomisation is performed continuously throughout the study following completion of the baseline assessment, without the use of a pre-generated allocation list. This procedure ensures allocation masking, prevents prediction of group assignment, and guarantees that each participant has an equal (50%) probability of being assigned to either study arm at the time of randomization. Participants begin the intervention within three working days from randomisation.

### Blinding

Clinical interviews and baseline parameters are conducted prior to randomisation, ensuring blinding for the participants during the study. Participants are informed that they will be randomised to one of two active digital interventions and are blinded to specific treatment allocation. The specific types of interventions remain undisclosed to them throughout and following the intervention. Due to the nature of the intervention, study staff are not blinded to participant allocation. Following study completion, a blinded statistician will be conducting the analysis of the results, which will be interpreted by the research group unblinded.

#### Data collection

Baseline interviews are performed by a cardiac study nurse and study psychologists. All screening and outcome questionnaires are administered via a secure web-based platform and server (BASS), provided by the eHealth Core Facility at Karolinska Institutet, Stockholm, Sweden. The questionnaires administration process is fully automated, with assessments automatically generated and reminders automatically sent to participants. Accelerometers (ActiGraph®) are distributed and returned via regular mail. This approach ensures a minimal of influence from study personnel on the data collection. Although, if a participant is late completing an assessment, study staff are notified, and the participant is contacted by study staff who remind the participant to complete the assessment. Participants are free to withdraw from the study at any time. In such cases, they will be asked to continue the data collection, i.e. completing self-assessments and wearing an accelerometer.

### Interventions

The CA-CBT and the CLI are administered over the internet through the BASS platform, which offers a secure delivery of digital behavioural interventions. Both intervention last eight weeks, and consist of eight interactive text-based modules in easy to read Swedish. The interventions are manualised, but build on an idiosyncratic and interactive approach. Participants have regular online contact with clinical psychologist and work through the modules in a step-wised manner, completing homework assignments each week. Participants submit their assignments (e.g. goals, behavioural checklists, exposure exercises, dietary or lifestyle behaviors) to their psychologist, and can ask questions via a message function in the platform. The psychologist guides the participant through the intervention and provide feedback on their assignments. The psychologist answers within two working days, via text-based communication in the messaging function in the platform.

Both interventions provide the same level of psychologist support and the psychologist guidelines in the setup are identical, except for the content of the interventions. In CA-CBT, the psychologist support focuses on exposure, while in CLI, it centers on psychological support. Both interventions include educational components on ACS. For the CA-CBT group, brief advice on lifestyle modifications following ACS is also included in the initial psychoeducation, to control for provision of general information and guidelines on lifestyle modification.

#### Training

The study psychologists include licensed psychologists and one psychologist in training, who is supervised by a licensed psychologist. In total, nine psychologists are expected to deliver the interventions, with all involved in both treatment arms. Prior to intervention delivery, all study psychologists receive training in CA-CBT and CLI, as well as structured education on ACS physiology and medical treatment, provided by study cardiologist and study cardiac nurse within the research team. They also receive training on the digital delivery of the interventions in the BASS4 platform and receive direct supervision in the platform from a licensed psychologist with expertise on CBT in cardiac patients. In addition to this, psychologists are able to consult and receive supervision in the platform from a cardiac study nurse and cardiologist (e.g. regarding case questions related to participants physical health), ensuring close multidisciplinary collaboration to prioritise participant safety.

#### CA-CBT

The CA-CBT has been tailored to the ACS-population and evaluated on patients experiencing cardiac anxiety following MI in previous studies (23) (Johnsson et al., manuscript in preparation; ClinicalTrials: NCT05580718). The CA-CBT intervention primarily targets cardiac anxiety in patients with ACS, focusing on: (1) fear of ACS recurrence; (2) fear, hypervigilance, or other affective arousal related to cardiac sensations; and (3) avoidance behaviours driven by fear, hypervigilance, or other emotional responses associated with the cardiac event. Common avoidance behaviours include being alone, or engaging in physical, emotional, or cognitive exertion (23).

The CA-CBT include the following components: *1. Education*: Psychoeducation on emotional reactions following ACS, primarily cardiac anxiety and typical behavioural adaptations. Education on ACS and lifestyle advice following ACS. Information on benign and acute cardiac symptoms, distinguishing between them and when to seek medical care. *2. Labeling:* Introduction and practice in labeling of bodily sensations, emotional responses, and behavioural impulses. Labeling involves taking a neutral and descriptive stance toward internal reactions, aiming to reduce the fear and emotional reactivity associated with bodily sensations, while also enhancing tolerance and the capacity to remain with these experiences until they gradually subside. *3. Interoceptive exposure:* Gradual exposure to cardiac-related and bodily sensations linked to cardiac anxiety by inducing similar bodily experiences, to reduce anxiety and increase tolerance to these sensations. The interoceptive exercises involve, for example, lying on the left side while focusing on cardiac sensations, inducing over breathing, and elevating heart rate through running in place. *4. In-vivo exposure:* A systematic, stepwise exposure to situations and activities that are associated with cardiac anxiety. The aim of the in-vivo exposure is to reduce limiting associations that interfere with daily functioning and restore QoL. The in-vivo exposure can involve situations such as visiting remote locations, planning for the future or participating in activities that are both engaging and physically demanding. It also incorporates a rationale for using exposure strategies to address common psychological symptoms post ACS such as general worry, depression, fatigue, fear of stress, and psychological reactions to pain. *5. Reduction of control and safety behaviours:* Actively and systematically reducing safety-seeking and control behaviours in anxiety-provoking situations, to decrease the reinforcement of anxiety.

Examples of response prevention include reducing behaviours such as pulse-checking, reassurance-seeking, and monitoring one’s whereabouts. *6. Relapse prevention:* Strategies to continue exposure and maintain treatment gains toward achieving goals. The relapse prevention includes planning for potential setbacks by identifying risk situations and proactively managing symptoms or stressors. The participants are also encouraged to combine the treatment strategies to enhance the effect of them, e.g., going for a walk on their own, while increasing their heart rate, and labeling bodily sensations and emotional responses. See Supplementary Material – Intervention Content for details on the distribution of components across the eight treatment modules.

#### CLI

The CLI includes education on lifestyle modifications and behavioural strategies to support adherence to general lifestyle recommendations following ACS. The content is based on guidelines on health promoting lifestyle modifications (26), which are offered as educational secondary prevention efforts in regular healthcare (27). The CLI also addresses common emotional reactions and psychological symptoms after ACS, such as stress and worry, and provides general guidance on the management of the psychological reactions, though it does not include a rational for the exposure-based approach. The CLI was created specifically for use as an attention control condition in this study and had not been evaluated prior to its deployment. The development of the intervention has been carried out with the involvement of cardiologist, cardiac nurses, physiotherapist and clinical psychologists.

The CLI contain the following components: *1. Education:* Information on ACS, treatments, common medical interventions and medication. Information on benign and acute cardiac symptoms, how to distinguishing between them and when to seek medical care. *2. Lifestyle advice:* Recommendations for lifestyle modifications, including adjustments in dietary habits, alcohol consumption, tobacco use, and physical activity. *3. Rationale on lifestyle recommendations and worry:* Psychoeducation on how ACS and subsequent lifestyle recommendations can, for some individuals, feel overwhelming and trigger uncertainty, stress, and worry. These reactions may arise due to perceived challenges, lack of support, or doubts about whether one is “doing it right,” potentially forming a vicious cycle of worry and uncertainty. The rationale emphasizes that worry about cardiac health can be improved when individuals receive appropriate support, relevant information, and guidance to focus on modifiable factors. With this support, patients can make informed and effective efforts, and thereby disrupt the cycle of worry and regaining a sense of control. *4. Behavioural strategies:* Introduction to general behavioral change strategies, including goal-setting, identifying barriers, stepwise progression, the use of rewards, and short and long-term consequenses. In addition, the CLI provides topic-specific behavioural change strategies, including guidance on reducing alcohol consumption, smoking cessation, and increasing physical activity. *5*.

*Common emotional reactions:* An overview of common emotional responses following ACS, such as stress, worry and depression, along with general psychological strategies for managing these reactions. The psychological strategies include validating emotional reactions, reducing unrealistic demands, and the encouraging a balanced approach between activity and recovery. *6. Relapse prevention:* Strategies aimed at sustaining lifestyle changes, prevent setbacks and relapse. See Supplementary Material – Intervention Content for a description of the CLI content in the intervention modules.

#### Safety considerations and multidisciplinary collaboration

There are no anticipated medical risks associated with participation in the trial. The content has been reviewed and approved by cardiologists, and previous studies on CA-CBT have reported no serious adverse events (23) (Johnsson et al., manuscript in preparation; ClinicalTrials: NCT05580718). Several essential safety measures are in place, including a pre-inclusion assessment by a cardiac nurse and cardiologist to ensure that participation poses no risk to the participants’ safety, and that the applicant have no contraindication to be physically active. The study psychologists also conduct inclusion assessments, and applicants with e.g. high-risk suicidal ideation are referred to standard care.

During the interventions, psychologists have access to on-demand supervision from a cardiac study nurse and cardiologist in the BASS platform, for instance, regarding participants’ reported symptoms or the adaptation of the exercises in the intervention. In case of symptoms that require medical attention, participants are referred to regular care and their treating physician. In addition to this, both intervention groups receive information on when to seek medical care and what constitutes benign and acute cardiac symptoms, e. g. benign palpitations during physical activity, and potentially more acute symptoms such as persistent, pressure-like, or cramping chest pain. Participants also self-report unwanted effects (28) weekly during the study, and at follow-up, to monitor adverse events.

### Outcomes

Demographic and clinical data are collected through participant self-reports and medical records. All self-reports in the trial are conducted digitally via the BASS platform.

#### Participant timeline and assessment points

All participants complete baseline assessments prior to randomisation. Weekly measurements are conducted throughout the eight-week intervention period, followed by post-intervention assessments and follow-ups at six and twelve months. Figure 2 illustrates the participant timeline and the specific timepoints at which outcomes are assessed.

**Figure 2.**
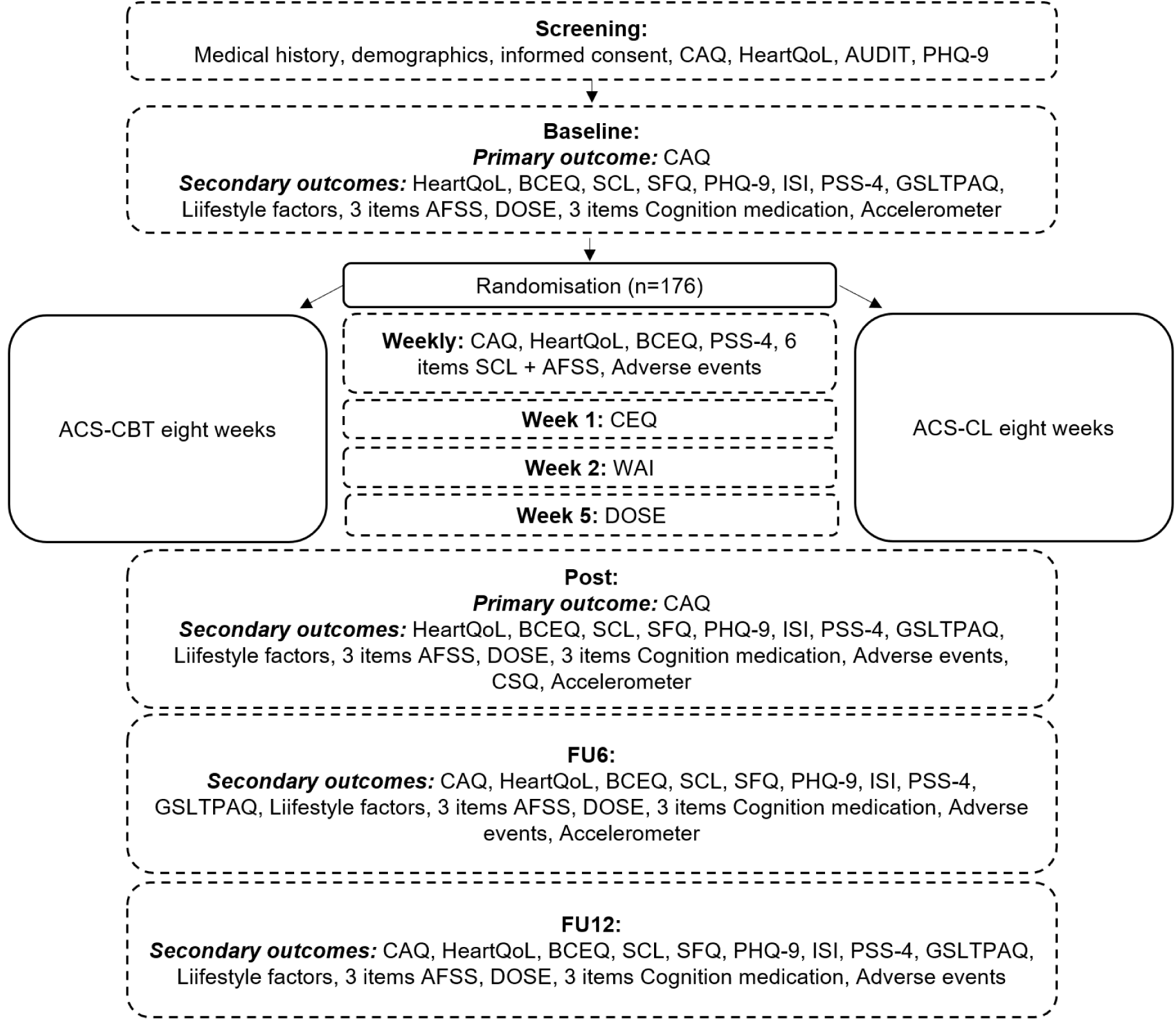
Participant timeline. CAQ, Cardiac Anxiety Questionnaire; HeartQoL, Heart Quality of Life; AUDIT, Alcohol Use Disorders Identification Test; PHQ-9, Patient Health Questionnaire-9; BCEQ, Behaviors following Cardiac Event Questionnaire; SCL, Symptom checklist Severity and Frequency Scale (adapted to coronary artery disease); SFQ, Short Fatigue questionnaire; ISI, Insomnia Severity Index; PSS-4, Perceived stress scale; GDLEQ, GDLEQ Leisure-time Exercise Questionnaire; IPAQ, International Physical Activity Questionnaire; AFSS, University of Toronto Atrial fibrillation Severity Scale; DOSE, Non-Adherence 3-item questionnaire; CEQ, Credibility/Expectancy Questionnaire; WAI-S, Working Alliance Inventory; CSQ-8, Client Satisfaction Questionnaire.

#### Primary outcome

The primary outcome is the mean difference in improvement in Cardiac anxiety questionnaire (CAQ) (8) score at post-intervention, based on the trajectories over the eight week intervention period. In total, the CAQ includes 18 items describing common behaviours and experiences related to cardiac anxiety, such as. “Even if the tests show normal results, I still worry about my heart”, “I avoid physical exertion” and “I can feel my heart in my chest”.

Participants rate the statements in terms of frequency on a five-point scale. In addition to the total score (primary outcome), the scale yields three subscale scores, fear/worry, avoidance, and attention–reflecting the different aspects of cardiac anxiety. Higher scores indicate greater levels of cardiac anxiety overall or within the specific subscale. The CAQ is well-validated and has demonstrated strong validity, e.g. in a Swedish population following myocardial infarction (29). The CAQ is collected at baseline, weekly during the interventions, post-intervention and at six– and 12-month follow-up.

#### Secondary outcomes

1. Disease-specific Health related QoL is assessed by Heart Quality of Life (HeartQoL). The scale comprises 14 items asking participants to rate how much their heart condition has bothered them, yielding a global score as well as physical and emotional subscales. Scores range from 0 to 3, with lower scores indicating lower QoL (30).
2. The Behaviors following Cardiac Event Questionnaire (BCEQ), developed by the research group, assesses cardiac-related avoidance behaviours through 17 statements, along with their frequency rated from “Never” to “Always” in five steps. Score range from 0-85.
3. Cardiac-related symptoms are assessed by Symptom checklist Severity and Frequency Scale (SCL, adapted to coronary artery disease) (31)
4. Fatigue is measured by the Short Fatigue questionnaire (SFQ), which assesses different aspects of fatigue through four statements. Participants are asked to rate the degree to which each statement is correct on a seven-point scale (32).
5. Depression levels are assessed using the Patient Health Questionnaire-9 (PHQ-9), which assess depressive symptoms and their impact on daily life through nine items (33).
6. Sleeping difficulties are measured using Insomnia Severity Index (ISI), in which participants are asked to rate the sleeping problems in seven questions (34).
7. The frequency of stress-related symptoms and levels of stress are assessed by the four item Perceived stress scale (PSS-4) (35).
8. To measure physical activity there are several outcomes: The Godin Leisure-time Exercise (GDLEQ) (36) assesses physical activity by measuring both the frequency and intensity of exercise. Additionally, physical inactivity is evaluated using a single item from the International Physical Activity Questionnaire (IPAQ)(37). Participants are also instructed to wear a wrist-worn accelerometer (ActiGraph®) continuously for seven days to measure physical activity levels, at baseline, post– and six-months following the intervention.
9. Modifiable risk factors are self-reported. This include demographic data, recent home or office blood-pressure and lifestyle factors. Lifestyle factors are reported through The Swedish national Board of health and Welfare questionnaire regarding diet (5 items), tobacco (2 items), alcohol (3 items), and BMI.
10. To measure cardiac-specific healthcare consumption three items regarding health-care seeking from the University of Toronto Atrial fibrillation Severity Scale (AFSS) (38) are distributed.
11. Medical adherence and prescription fill-rate are assessed through the DOSE Non-Adherence 3-item questionnaire and 1-item on non-adherence with regards to side effects (39). In addition to this, the prescription fill-rate of cardiovascular medication (e.g., statin) from the Swedish Prescribed Drug Register (40) are collected with a retrospective period of six months.
12. Aversive cognition toward medication is measured using a three-item subset of questions addressing psychological distress related to cardiac medication (41).

#### Weekly outcomes

To maximise statistical power, monitor participants over the treatment period and in preparation for a mediation analysis, the following measures are assessed weekly during the treatment period: cardiac anxiety; CAQ (8), disease specific QoL; HeartQoL(30), behaviours related to cardiac anxiety; BCEQ five items and perceived stress; PSS-4 (35). In addition to this, four items measuring cardiac-related symptoms from SCL (31) and two items from the University of Toronto Atrial fibrillation Severity Scale (AFSS) (38) are administrated weekly.

#### Process outcomes

1. Therapeutic factors such as treatment credibility and alliance are measured through two questionnaires: The Credibility/Expectancy Questionnaire (CEQ), which assesses participants’ perceptions of treatment credibility and their expectations of its effectiveness using five questions (42). The Working Alliance Inventory (WAI-S) measures the therapeutic alliance using 12 questions (43). Higher scores on both questionnaires reflect more positive outcomes, indicating greater treatment credibility and a stronger therapeutic alliance, respectively.
2. Adverse effects of the treatment are captured using a questionnaire in which participants report adverse events and the level of impact on the participant (28). Satisfaction with the treatment is assessed by the Client Satisfaction Questionnaire (CSQ-8) which measures contentment using eight questions, with higher sum scores indicating greater satisfaction (44).

### Sample size and power

The sample size in the study is based on data obtained from our RCT of digital CBT for cardiac anxiety following MI. Data for weekly measures during treatment were extracted for the CA-CBT group and analyzed by an independent statistician. Within– or between-group effects were not calculated, but instead the power analysis was based on the observed within– and between-individual variance on the weekly measures and on a pre-determined threshold of relative efficacy of CLI compared to CA-CBT. We assumed that the effect of CA-CBT in the present trial will be similar to the previous RCT and that the CLI group will show 85% or less of the effect of CA-CBT. This power analyses yielded a sample size of 75 patients per group for 80% power at alpha level 0.05. Allowing for a 15% dropout, we aim to include 88 patients per group, 176 in total.

### Statistical methods

Analyses will be conducted according to the “intention-to-treat” principle, including all randomised participants. A linear mixed-effects model with random intercepts will be used to analyse the primary outcome–the difference in average change on CAQ scores over the tenweek measurement period (baseline, eight weekly assessments during treatment, and post-intervention) between the CA-CBT and CLI groups – with time, treatment, the interaction effect between time and treatment as independent variables, and a random intercept. Effect sizes will be calculated as the estimated standardised mean difference at post-intervention (Cohen’s d) based on the baseline standard deviation, and the alpha level is set at .005.

Most of the secondary outcomes will be analysed using similar methods as for the primary outcome but including fewer time-points (baseline, post, and six-month follow-up). In addition to this, we will also analyse the secondary outcomes dichotomised and descriptively within categories using logistic mixed-effects model. This will allow us to investigate the proportion of participants who transition from one category to another, e.g., from below the recommended levels of physical activity to within the recommended levels.

Process outcomes are collected once, and the groups will be compared with t-test regarding these outcomes. Adverse events will be analysed by comparing the number of reported events, and the rated severity of the events.

#### Data management and monitoring

All self-reported data are collected in the secure web-based platform (BASS) provided by the eHealth Core Facility at Karolinska Institutet. The system ensures secure data entry, storage, management and confidentiality. Automated processes are in place for questionnaire distribution and reminders, and data quality is promoted through structured input formats and regular monitoring by study staff. No interim analysis will be conducted, and no data monitoring committee will be formed, as we anticipate a rapid recruitment pace. Additionally, no increased risks are expected from participation in the study. No formal auditing of trial conduct is planned, and a data monitoring committee is not considered necessary for this study, due to the low-risk nature of the digital, non-invasive interventions and the presence of regular safety and procedural monitoring within the research team.

### Patient and public involvement

The CA-CBT was developed and evaluated in three previous studies (23) (Johnsson et al., manuscript in preparation; ClinicalTrials: NCT05580718). The development included the integration of participants experiences and feedback from the previous studies. Adaptations drawn from participant feedback included the addition of clinical vignettes, for different clinical displays of cardiac anxiety, and examples on how the participants with a specific type of clinical display worked with the assignments in the intervention. The adaptation also included the shortening of the intervention texts, as they were deemed as too long by participants. Additionally, the protocol and measurement tools were adjusted between the studies to better align with the target population.

### Clinical implications

This research project has the potential to integrate digital CBT for cardiac anxiety following ACS as a conjunction to current treatment options, such as cardiac rehabilitation. The integration could promote a multidisciplinary approach that incorporates psychological evaluation and treatment into ACS aftercare. If the CA-CBT continues to demonstrate efficacy it may thus constitute an important addition to current treatment options and clinical care, with potential for accessibility and implementation in clinical practice to benefit the target population.

### Ethics and dissemination

#### Ethics

All gathered patient information are protected according to Swedish regulations, and performed in accordance with the Declaration of Helsinki. The trial began following approval from the Swedish Ethical Review Authority; Dnr 2023-07605-01, and is registered at ClinicalTrials; NCT06298864. The reports will be prepared according to the CONSORT-guidelines for non-pharmacological treatment studies. Any changes to the protocol will be communicated via ClinicalTrials.gov and reported to the Swedish Ethical Review Authority in accordance with applicable regulations. Before inclusion, each patient receives both oral and written information on what participation in the study include, and give informed consent to participation. All participants are informed that they may withdraw from the study whenever they wish, and that they in case of withdrawal are treated according to the best possible routine standards. The informed consent form is available as Supplementary Material.

#### Harms

To reduce harm there are several aspects that we have considered. First, there have been no serious risks associated with our previous studies on the CA-CBT. Although, mild and transient side effects, such as stress during the intervention and increased anxiety during exposure, were reported. Importantly, the unwanted effects were temporary and passing (23) (Johnsson et al., manuscript in preparation, ClinicalTrials: NCT05580718). Second, interventions focusing on lifestyle modification (like the CLI) are recommended in guidelines (12), and the CLI does not scope beyond the traditional advice in such interventions. Third, participation in either the CA-CBT or CLI may offer potential health benefits, including a potential reduction in anxiety and/or improved health behaviours, with higher adherence to lifestyle recommendations. Forth, adverse events will be collected weekly during the trial, and at follow-up measurements to minimise harm. And fifth, the inclusion criteria of ACS ≥ 6 months before assessment helps ensure that participation in the trial does not interfere with the recovery or aftercare following the cardiac event. Thus, no harm is expected as a result of participation in the trial.

#### Trial status

The study started recruitment in March of 2024 and the first patient was included into the trial on March 24 2024. As of June 5, 2025, 81 patients have been randomised in the trial. Recruitment will continue until a total of 176 participants are, which is expected to be achieved in the latter half of 2026.

#### Dissemination

The results from the trial will be published upon study completion and submitted to a peer-reviewed scientific journal. Authorship follows the guidelines of the International Committee of Medical Journal Editors, with the order determined by each authors level of involvement in the study. The individual participant data underlying the analyses in this study cannot be made publicly available due to Swedish and European Union data protection regulations. Requests for additional outcomes or estimates may be directed to the corresponding author and will be handled in accordance with legal expertise and the sponsor’s current data governance guidelines.

## Funding

This study was supported by the Swedish Heart and Lung foundation and the Swedish research Council. The funding body was not involved in the design of the study, data analysis, or interpretation of the results.

## Conflict of interest disclosures

Dr Braunschweig declares personal fees for trial committee participation and lectures by Medtronic, Biotronik, Biosense Webster, Impulse Dynamics, Novartis, Orion, Boehringer and Pfizer. Dr Ljótsson has co-authored a Swedish self-help book on exposure-based cognitive behaviour therapy for health anxiety and has a publishing agreement with Cambridge University Press for a self-help book for irritable bowel syndrome. Dr Mellbin declares lectures, consulting and clinical trials fees to her institution from Amarin, Amgen, Astra Zeneca, Bayer AG, Boehringer-Ingelheim Janssen, Novartis, NovoNordisk, Sanofi. No other disclosures were reported.

## Protocol version

1.0 (Date: 6 June 2025)

## Supporting information

SPIRIT checklist

Supplementary Material – Intervention Content

informed consent form Supplementary material

## Data Availability

The individual participant data underlying the analyses in this study cannot be made publicly available due to Swedish and European Union data protection regulations. Requests for additional outcomes or estimates may be directed to the corresponding author and will be handled in accordance with legal expertise and the sponsors current data governance guidelines.

## Abbreviations

CBT: Cognitive behavioural therapy
ACS: Acute coronary syndromes
CA-CBT: Cognitive behavioural therapy targeting Cardiac Anxiety
RCT: Randomized controlled trial
QoL: Quality of Life

## Contributors

Amanda Johnsson contributed to study conceptualisation, treatment development and adaptation, data collection, supervision as a psychologist, treatment delivery, and drafting and revising the manuscript. Brjánn Ljótsson contributed to study conceptualisation, manuscript writing and revision, and secured resources and funding. Frieder Braunschweig contributed to study conceptualization, reviewed and edited the manuscript. Linda G Mellbin served as the supervising study cardiologist, contributed to study conceptualisation, and reviewed and edited the manuscript. Josefin Särnholm contributed to study conceptualisation, treatment development and adaptation, supervision as a psychologist, and drafting and revising the manuscript.

## Notes

### Clinical Trial

NCT05580718

### Author Declarations

Ethical Review Authority of Sweden gave ethical approval for this work, Dnr 2023-07605-01

