## Supplementary Material – Intervention Content for "Digital Cognitive Behavioral Therapy for Cardiac Anxiety Following Acute Coronary Syndrome: Protocol for a Randomised Controlled Trial Comparing CBT to a Digital Lifestyle Intervention"

**Supplementary Textbox. 1. Contents of CA-CBT**

| | |
|---|---|
| Module 1 - Introduction and psychoeducation | • Common emotional responses following ACS<br>• The relationship between cardiac anxiety and avoidance behavior<br>• Identifying avoidance and control behaviours<br>• Goal-setting<br>• Labeling Education on ACS, risk factors, its treatments and medication |
| Module 2 - Interoceptive exposure | • Exposure to physical sensations to reduce the associated fear<br>• General lifestyle advice on e.g. physical activity |
| Module 3 - Introduction to exposure in-vivo | • Gradual exposure to avoided situations, places, and activities<br>• Reduction of avoidance and control behaviours |
| Module 4 – Continuing exposure in more areas | • Rationale for exposure strategies to manage , e.g. worry, depression, fatigue, stress, and pain |
| Modules 5, 6, 7 - Continuing exposure and reclaiming activities | • Continuous work with gradual interoceptive and in-vivo exposure |

| | |
|---|---|
| | • Combining the treatment strategies |
| Module 8 - Summary and relapse prevention | • Summary of treatment<br>• Identifying risk situations<br>• Plan for future work towards goals |

**Supplementary Textbox 2. Contents of CLI**

| | |
|---|---|
| Module 1 – About ACS, causes and treatment | • Education on ACS, risk factors, its treatments and medication<br>• Rationale on how support in implementing lifestyle changes after ACS can reduce health-related worry and promote a sense of control.<br>• Behavioural change strategies targeting health behaviours, such as goal setting, rewards, identifying barriers, stepwise progression, identifying short and long-term consequenses |
| Module 2 - Dietary habits, alcohol and tobacco | • Education and advice promoting healthy habits regarding diet, alcohol and tobacco<br>• Behavioural strategies related to the areas |

| | |
|---|---|
| Module 3 - Physical activity | • Education regarding physical activity and the beneficial effects on health<br>• Behavioural strategies related to physical activity |
| Module 4 - Common emotional reactions | • Education regarding common emotional reactions following ACS<br>• Tools related to emotional reactions such as…. |
| Module 5, 6, 7 – Continued work with relevant areas | • Repetition of the relevant information and behavioural strategies from previous modules |
| Module 8 - Maintain a healthy lifestyle | • Prevention of relapse and plan forward to maintain a healthy lifestyle |
