## Supplementary material for "Digital Cognitive Behavioral Therapy for Cardiac Anxiety Following Acute Coronary Syndrome: Protocol for a Randomised Controlled Trial Comparing CBT to a Digital Lifestyle Intervention": informed consent form Supplementary material

### Participant Information: Invitation to Participate in a Study on Internet-Based Psychological Support After Myocardial Infarction or Unstable Angina

#### Background and Purpose

You are invited to participate in a study comparing two internet-based programs following myocardial infarction or unstable angina. This document provides information about the study and what participation entails.

We aim to investigate whether internet-based psychological support can improve quality of life, mental well-being, and physical activity in individuals who have experienced a myocardial infarction or unstable angina. We also want to explore experiences related to taking heart medications and patterns of medication use.

Myocardial infarction and unstable angina are stressful life events that can affect both physical and mental health. Research shows that many patients experience depression, anxiety, fatigue, and reduced quality of life, and that taking heart medications can be perceived as burdensome. Concerns about heart health can negatively impact daily life.

Because these symptoms and psychological reactions are common and often distressing, we have developed two internet-based programs tailored to address these issues. Internet-based psychological support is well-established and has proven effective in other studies involving physical illnesses that affect quality of life and well-being.

The purpose of this study is to evaluate and compare two types of internet-based programs specifically designed for individuals who have had a myocardial infarction or unstable angina. We want to assess whether these programs can improve quality of life and reduce distress, as well as examine medication use and experiences related to taking heart medications.

#### Who Can Participate?

This study is intended for individuals who had a myocardial infarction or unstable angina more than six months ago and who experience worry about their heart and heart health that affects their daily life. Internet-based psychological support may help you feel better and gain greater control over your life.

To determine eligibility, we will first conduct a medical and psychological assessment.

#### How to Register and Participate in the Study

Participation involves several steps:

**Step 1: Read the Information.** Please read this entire document. If you wish to express interest in participating, you consent to the study as described here. You may also contact the study coordinator by phone or email (see contact details below) if you have questions.

By consenting to participate, you also agree that we may access your previous medical records related to your heart condition. You also consent to the storage of your personal data.

**Step 2: Online Questionnaires.** If you consent, you will be directed to a secure website where you will complete questionnaires about your health, heart-related anxiety, mood, and alcohol habits. This takes about 15 minutes.

**Step 3: Nurse Interview.** After we receive your registration, a nurse will call you for a telephone interview. You will be asked about your heart event, current heart health, blood pressure, and medications. The purpose is to assess whether participation is suitable for you. You will also have the opportunity to ask questions and confirm your interest in participating.

**Step 4: Psychologist Interview.** If no obstacles are found, you will be contacted by a psychologist for an interview. The psychologist will ask questions to help determine whether the internet-based programs are appropriate for you. You will also receive information about how the programs work and can ask further questions.

**Step 5: Cardiologist Review.** If participation still seems appropriate, a cardiologist will review the overall assessment before a final decision is made. If participation is deemed unsuitable, you will be informed and referred to regular healthcare if needed.

*You may withdraw from the study at any time without giving a reason. We may ask why, but you are n Measurements and Follow-Up*

#### Pre-Assessment

Before starting either of the internet-based programs, we want to assess your quality of life, mental well-being, and physical activity. This is done by completing online questionnaires, which take about 30–40 minutes. You will also wear an activity watch on your wrist for one week. Additionally, we will ask you to measure your blood pressure.

#### Randomisation

After completing the pre-assessments, you will be randomly assigned to one of the two internet-based programs, each lasting 8 weeks.

#### Internet-Based Psychological Support

In the programs, you will explore and receive strategies for changing behavioral patterns and lifestyle habits that may help reduce symptoms after a cardiac event and improve quality of life. You will have access to written material and work through one chapter with exercises each week. The exercises are safe and have been developed in consultation with cardiologists.

Each week, you will have contact with a psychologist via the internet. The psychologist will support you in working through the program, provide advice and answer questions. The program does not require more computer skills than what is needed to send a regular email.

Participation requires a high level of personal responsibility and engagement. It is recommended that you set aside at least 30 minutes per day for exercises and homework. The program runs for 8 weeks. Short breaks (e.g., for holidays) are allowed, but if you anticipate difficulty committing time during this period, you should not participate.

#### Assessments and Follow-up

**Questionnaires.** Before starting the program, you will complete the same questionnaires mentioned in the pre-assessment. You will also fill out short weekly questionnaires during the program, which take about 10 minutes per week. After completing the program, and again at 6 and 12 months, you will fill out the same questionnaires once more. Your responses are important for evaluating the effects of the programs.

**Activity Watch.** All participants will wear an activity watch before, immediately after, and 6 months after the treatment. The watch is worn on the wrist for one week and measures your physical activity.

**Blood Pressure.** You will be asked to measure your blood pressure before, after, and 6 months after the program. This can be done at home or at your healthcare center. ot required to explain.

**Heart Medications.** Before starting the program, and again at 6 and 12 months after completion, we will extract data from the national prescription register to analyze heart medication usage at the group level. At each time point, we will review prescriptions and dispensations over the previous 6 months.

#### Other Psychological or Medical Treatments

You should avoid starting other psychological treatments or making significant medication changes or undergoing procedures during the 8-week program. However, your current physician remains responsible for your medical care and will make decisions together with you. Please inform us if you plan to begin any of the above during the program.

#### Benefits and Risks of Participation

Previous studies on internet-based support for heart disease and other conditions suggest that following the program's guidance may improve your quality of life and reduce current symptoms. To our knowledge, the exercises included pose no health risks. All exercises and advice have been approved by cardiologists.

However, since the programs address common reactions and symptoms after a cardiac event, you may experience increased focus on your heart and related symptoms during the program, which can be distressing. This is usually temporary. The program also requires a significant time commitment, which may feel stressful.

Your psychologist will support and guide you through the program and help you plan your exercises to ensure you benefit from them, even if they cause some short-term discomfort. If needed, the psychologist will also consult with the study's cardiologist regarding your progress.

#### What Happens to My Data?

The project will collect and register information about you. As a participant in the study, you will be asked to provide personal data such as your name, personal identity number, gender, education, and marital status, as well as information about your social situation and health. It is your decision whether to provide this information, but we require it in order for you to participate in the study. Your data will be processed for the scientific purpose of evaluating the effects of internet-based programs following myocardial infarction or unstable angina, and to assess the functioning of the questionnaires you complete.

Your personal data will be processed in accordance with the EU General Data Protection Regulation (GDPR) for the purpose of research, based on the legal ground of public interest. Your responses and results will be stored in secure, classified databases at Karolinska Institutet (KI). Traffic to and from our digital platform is encrypted, and login requires a personal account with two-factor authentication. Sensitive information is handled confidentially.

During the internet-based program, your psychologist may access your data in order to provide you with the best possible care. Otherwise, the information will only be handled by researchers and research assistants involved in the study, and your personal data will never be disclosed to unauthorized individuals. We process your information in pseudonymized form, meaning without sensitive identifiers such as your name or personal identity number. Your responses and results will be handled in a way that prevents unauthorized access.

Data from the study will be archived in accordance with KI's current guidelines. After a maximum of 10 years, the code key will be destroyed, after which it will no longer be possible to retrieve any register extracts. All project staff are bound by healthcare confidentiality regulations. All results from the study will be reported in a way that ensures you cannot be identified as an individual participant.

Your responses and results will be handled in such a way that unauthorized individuals cannot access them. The data controller responsible for your personal data is Karolinska University Hospital, Region Stockholm. According to the EU General Data Protection Regulation (GDPR), you have the right to access, free of charge, the personal data about you that is processed in the project, and to request correction of any inaccuracies. You may also request that your data be deleted or that the processing of your personal data be restricted. However, the right to deletion and restriction does not apply if the data is necessary for the ongoing research. If you wish to access your data, please contact: Josefin Särnholm, +46 (0)8-524 832 58 Data Protection Officer:. If you are dissatisfied with how your personal data is being processed, you have the right to file a complaint with the Swedish Authority for Privacy Protection (Integritetsskyddsmyndigheten), which is the supervisory authority.

#### Study Results

After the 12-month follow-up, we will compile the results of the programs. When you complete the 12-month follow-up, you will have the opportunity to indicate whether you wish to receive the results.

#### Insurance and Compensation

Participation is free of charge. No financial compensation is provided. Standard patient injury insurance applies to this study.

#### Voluntary Participation

Participation is voluntary. You may withdraw at any time without giving a reason. If you choose not to participate or to withdraw, it will not affect your future care or treatment. If you withdraw, we may ask if you are willing to complete a few questionnaires online or explain your reasons by phone—this is entirely voluntary.

To withdraw, please contact the project lead (see below).

#### Responsible Parties

This research project is a collaboration between Karolinska University Hospital and Karolinska Institutet. Karolinska University Hospital is the principal investigator.

Principal Investigator:

Josefin Särnholm, Licensed Psychologist, PhD

Department of Clinical Neuroscience, Division of Psychology

Karolinska Institutet

Nobels väg 9, 171 65 Stockholm

 | 08-524 832 58

Lead Cardiologist and Medical Supervisor:

Linda Mellbin, Senior Consultant Cardiologist, Associate Professor

Department of Cardiology, Karolinska University Hospital

Study Contact Person:

Amanda Johnsson, Licensed Psychologist

Department of Clinical Neuroscience, Division of Psychology

Karolinska Institutet

Nobels väg 9, 171 65 Stockholm

 | 08-524 824 50

#### Consent to Participate in the Study

To complete your registration, you must confirm and agree to the following. If you cannot agree, you may close your browser, and your registration will not be processed.

Before registering, you must have read the full participant information.

☐ I confirm that I have read the entire participant information and consent to participate in the study: *Internet-Based Behavioral Intervention with Psychological Support After Myocardial Infarction or Unstable Angina – A Randomized Controlled Trial*

☐ I consent to the research team accessing my medical records related to my heart health.

☐ I consent to the extraction of data regarding my use of heart-related medications from the Swedish Prescribed Drug Register, as described in the participant information.

☐ I consent to the processing of my personal data as described in the participant information.

We may need to contact you via email or SMS to schedule assessment interviews. After the treatment, we may also contact you via email or SMS to remind you to complete follow-up assessments.

☐ I consent to being contacted via email or SMS for scheduling assessment interviews and reminders about follow-up assessments.

☐ I understand that I may withdraw from the study at any time without needing to provide a reason, and that doing so will not affect my access to other forms of care.
